# Analytical Study of the Medical Management of Pregnancy Loss: Implementation of the FIGO Misoprostol Protocol

**DOI:** 10.64898/2026.01.16.26344127

**Authors:** Boumaiza Amel, Tharwa Necib, Garci Meriem, Mahdi Makni, Ameni Abdeljabbar, Abir Chaouachi, Olfa Slimani, Nabil Mathlouthi, Cyrine Belghith

## Abstract

Medical management of pregnancy loss and termination has evolved considerably over the past decade, with misoprostol-based protocols emerging as a cornerstone of care in many settings. Standardized regimens, such as the FIGO 2017 recommendations, aim to improve outcomes by ensuring effectiveness, safety, and reproducibility across diverse clinical contexts. Evaluating their real-world performance is essential, particularly in resource-constrained environments where protocol adherence can directly influence patient care.

**Objective:** To evaluate the effectiveness, safety, and clinical applicability of the FIGO 2017 misoprostol protocol for medical management of first□trimester missed abortions, second□trimester intrauterine fetal demise (IUFD), and medical termination of pregnancy (MTP) in a Tunisian tertiary center.

**Methods:** A retrospective descriptive and analytical study was conducted in the Gynecology Department A of Charles Nicole Hospital between January 2022 and December 2023. All patients admitted for first□trimester missed miscarriage, IUFD, or MTP and managed with misoprostol were included.

**Results:** A total of 190 patients were included: 65.8% with first□trimester missed abortion, 14.4% with IUFD, and 19.8% with MTP. The global success rate was 89.7%. Median induction□to□expulsion time was 32 h for first□trimester pregnancy loss, 19 h for IUFD, and 21 h for MTP. Multigravida women experienced significantly shorter expulsion intervals. Complications occurred in 8.8% of cases, with no uterine rupture.

**Conclusion:** The FIGO 2017 misoprostol protocol is highly effective and safe in the Tunisian context, offering high success rates, shorter hospitalization, and minimal complications. Findings support its continued implementation and highlight the need for further research in women with uterine scars.

## 1. Introduction

Early pregnancy loss, intrauterine fetal demise (IUFD), and medically indicated termination of pregnancy represent common yet complex clinical situations requiring effective and compassionate management. Misoprostol, a synthetic prostaglandin E1 analogue, is widely used due to its efficacy, low cost, and stability at room temperature. The International Federation of Gynecology and Obstetrics (FIGO) updated its recommendations in 2017 to optimize dosage, route, and administration intervals.

In Tunisia, misoprostol has become standard practice for medical evacuation, but real□world data evaluating protocol performance remain limited. This study assesses the effectiveness, safety, and clinical applicability of the FIGO 2017 protocol in a tertiary hospital.

## 2. Methods

This analytical and descriptive study was carried out over two years (January 2022 – December 2023) in the Gynecology Department A of Charles Nicole Hospital. All patients admitted for first-trimester missed abortion, IUFD, or medically indicated pregnancy termination and treated with misoprostol according to the FIGO 2017 protocol were included. Sociodemographic characteristics, obstetric history, misoprostol regimen, induction-expulsion interval, success rates, and complications were collected and analyzed using SPSS 25.*

## 3. Results

A total of cases meeting the inclusion criteria were analyzed. Pregnancy outcomes varied across maternal age groups (Table 1). Missed abortions were most frequent among women aged 30– 39 years (40.7%), followed by those aged 20–29 years (34%). IUFD was also more common in the 30–39-year group (47.6%). Medically indicated terminations (MTP) showed a similar age distribution, with 41.4% occurring in women aged 30–39 years.

**Table 1:** Pregnancy Outcomes by Maternal Age Goup.

| **Maternal Age(Years)** | **Missed Abortion (%)** | **IUFD (%)** | **MTP (%)** |
| --- | --- | --- | --- |
| **17-19** | **7.8** | **0** | **6.9** |
| **20-29** | **34** | **33.3** | **34.5** |
| **30-39** | **40.7** | **47.6** | **41.4** |
| **>40** | **11.6** | **19.5** | **15.1** |

Regarding induction-to-expulsion interval (Table 2), IUFD cases showed the shortest mean duration (19 h) and the highest success rate at 24 hours (90.4%). The mean interval was longer in missed abortions (32 h), with a 24-hour success rate of 56.7%. For MTP, the mean induction-to-expulsion interval was 21 hours, with a success rate of 67% at 24 hours.

**Table 2:** Induction to Epulsion Interval by Indication :

| **Indication** | **Mean Interval (h)** | **Success at 24 h (%)** |
| --- | --- | --- |
| **Missed abortion** | **32** | **56.7** |
| **IUFD** | **19** | **90.4** |
| **MTP** | **21** | **67** |

Success rates according to misoprostol regimen are detailed in Table 3. In missed abortions, vaginal administration of 800 µg every 3 hours resulted in a 24-hour success rate of 56.7% and an overall success rate of 79.4%. The sublingual 600 µg regimen demonstrated slightly higher success rates (62.3% at 24 hours and 80.1% overall). For IUFD, both regimens (100– 200 µg VV every 6 hours) achieved excellent results, with overall success rates of 100%. In MTP, success rates ranged from 67% to 73.2% at 24 hours, with total success close to 90%.

**Table 3:** Success Rates According To Misoprostol Regimen.

| Indication | Regimen | Success at 24 h (%) | Total Success (%) |
| --- | --- | --- | --- |
| Missed abortion | 800 µg VV / 3h | 56.7 | 79.4 |
| Missed abortion | 600 µg SL / 3h | 62.3 | 80.1 |
| IUFD | 200 µg VV / 6h | 90.4 | 100 |
| IUFD | 100 µg VV / 6h | 92.5 | 100 |

Complications are summarized in Table 4. Retained products were the most common complication (5.2%). Severe hemorrhage and infection were rare (0.5% and 1%, respectively). No cases of uterine rupture were recorded.

**Table 4:** Complications And Adverse Effects :

| Complication | Frequency (n) | Percentage (%) |
| --- | --- | --- |
| Retained products | 10 | 5.2 |
| Severe hemorrhage | 1 | 0.5 |
| Infection | 2 | 1.0 |
| Uterine rupture | 0 | 0 |

## DISCUSSION

The management of early pregnancy loss, intrauterine fetal demise (IUFD), and medical termination of pregnancy (TOP) remains a significant challenge in obstetric practice, requiring therapeutic strategies that are both effective and respectful of women’s physical and emotional well-being. Misoprostol, as standardized in the 2017 FIGO recommendations, has emerged as a cornerstone option due to its efficacy, safety, and ease of use (1–3).

In this study, the overall success rate of misoprostol reached 89.7%, closely mirroring international evidence that consistently reports success rates exceeding 80% when misoprostol alone is used for medical management (4–6). These findings reinforce the drug’s reliability across different clinical scenarios, particularly when protocols are carefully followed. The induction-to-expulsion intervals observed—especially the shorter duration in second-trimester IUFD—fit within the ranges documented in the literature, where expulsions typically occur between 12 and 36 hours depending on gestational age, uterine sensitivity, and dosing regimen (7,8).

In first-trimester pregnancy loss, the sublingual regimen demonstrated a slightly higher efficacy and faster action compared to the vaginal route in our cohort. This trend is well supported by pharmacokinetic studies showing faster absorption, higher peak serum levels, and greater bioavailability of sublingual misoprostol, which likely translates into shorter induction intervals (9,10). Nevertheless, both routes remain validated by FIGO and WHO, offering valuable flexibility in clinical practice.

The management of women with previous cesarean delivery is an area of continued caution. Although no uterine rupture occurred in our sample, the literature consistently notes an increased risk of rupture when higher doses of misoprostol are used in scarred uteri (11–13). FIGO recommendations for dose reduction in such women are therefore strongly justified, and the reassuring absence of complications in our cohort likely reflects both the relatively small number of scarred uteri and strict adherence to the protocol. Future studies with larger subgroups may help refine dose adjustments and better quantify risk.

Complications in our study were infrequent and generally mild, echoing rates reported in major clinical trials (14–16). Retained products of conception were the most common issue, but their frequency remained low. Severe hemorrhage and infection were rare, and the absence of uterine rupture confirms misoprostol’s strong safety profile in a controlled setting. Such outcomes highlight the importance of structured supervision, patient counseling, and timely follow-up, especially in low-resource environments where misoprostol is often the most accessible option.

Beyond its pharmacological dimensions, the emotional burden associated with pregnancy loss warrants equal attention. Several studies emphasize the psychological distress experienced by women undergoing IUFD or TOP, advocating systematic psychological support as part of comprehensive care (17). Although psychological outcomes were not specifically assessed in our study, integrating mental health support could represent an important improvement for patient-centered care in Tunisian hospitals.

Overall, our findings align closely with WHO, ACOG, and FIGO guidelines. They reaffirm misoprostol’s position as a safe, effective, and adaptable drug for managing early pregnancy loss and second-trimester fetal demise (1–3,18–20). The consistency between our results and international evidence strengthens the case for maintaining the FIGO 2017 protocol as the standard of care in Tunisia and similar settings.

## Conclusion

Missed abortion and second-trimester IUFD are frequent and emotionally taxing situations that require management strategies balancing efficiency, safety, and compassion. The application of the FIGO 2017 misoprostol protocol in our center demonstrated high success rates, short induction-to-expulsion intervals, and a low complication profile. These results confirm the protocol’s suitability for routine use and its capacity to reduce reliance on surgical procedures, particularly in resource-limited contexts.

In Tunisia, the implementation of standardized misoprostol regimens has proven effective across a wide range of obstetric scenarios. Continued adherence to international guidelines, coupled with tailored adjustments for women with uterine scars, will further enhance patient safety. Future research should focus on refining dosing strategies in scarred uteri, evaluating patient satisfaction, and exploring long-term psychological and reproductive outcomes.

## Data Availability

All data produced in the present work are contained in the manuscript

